# Transportability of patient outcomes from a US clinical trial to real-world populations - a case study using Lung-MAP S1400I (NCT02785952)

**DOI:** 10.1101/2024.05.25.24307916

**Authors:** Alind Gupta, Kelvin Chan, Manuel Gomes, Stephen Duffield, Sreeram Ramagopalan, Seamus Kent, Vivek Subbiah, Winson Cheung, Eran Bendavid, Paul Arora

## Abstract

2.

**Background:** The external validity of results from clinical trials to routine clinical practice is often questioned. This is sometimes because certain real world patient groups are excluded or underrepresented in clinical trials, or because standards of care in trials are different from those in real-world populations globally. This lack of external validity of trial results manifests as an efficacy-effectiveness gap. In this study, we aim to address the question of whether it is possible to extend results from a clinical trial to real-world populations across different countries. To do this, we use the Lung-MAP nonmatch sub-study S1400I trial (NCT02785952) as a case study.

**Setting:** Squamous cell lung carcinoma is a subtype of non-small cell lung cancer (NSCLC) accounting for 25-30% of cases. Compared to other NSCLC subtypes such as adenocarcinoma, the presence of actionable genetic variants is less common and there are fewer targeted therapies available for advanced/metastatic NSCLC (aNSCLC) of squamous subtype. Patients with squamous aNSCLC who progress on front-line chemotherapy commonly receive immunotherapy using immune checkpoint inhibitors such as nivolumab. The Lung-MAP nonmatch sub-study S1400I (NCT02785952) compared overall survival (OS) in patients with recurrent/stage IV squamous NSCLC randomized to receive either nivolumab monotherapy or nivolumab + ipilimumab combination therapy and found no significant difference in mortality rates between these groups. The trial included patients from the United States only.

**Objectives:** The goal of this study is to evaluate the transportability of results from NCT02785952 in United States patients to real-world populations in the United States, Germany, France, England and Japan. Using individual-level data for OS from NCT02785952, we will adjust for baseline characteristics from published studies of real-world populations in these countries and benchmark the predicted OS against Kaplan-Meier estimates reported by these studies for patients with squamous cell aNSCLC treated with nivolumab. Sensitivity analyses for unmeasured prognostic variables will be performed.

## 3 Amendments and updates

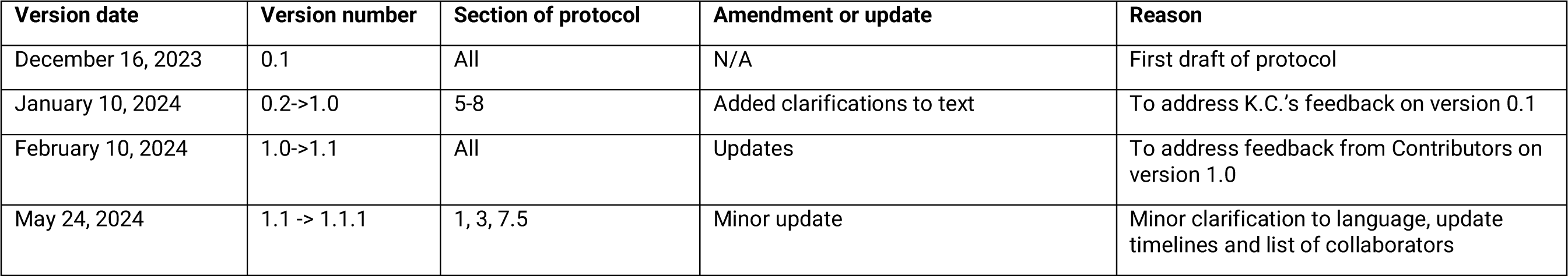

3. Amendments and updates
| Version date | Version number | Section of protocol | Amendment or update | Reason |
| --- | --- | --- | --- | --- |
| December 16, 2023 | 0.1 | All | N/A | First draft of protocol |
| January 10, 2024 | 0.2->1.0 | 5-8 | Added clarifications to text | To address K.C.'s feedback on version 0.1 |
| February 10, 2024 | 1.0->1.1 | All | Updates | To address feedback from Contributors on version 1.0 |
| May 24, 2024 | 1.1 -> 1.1.1 | 1, 3, 7.5 | Minor update | Minor clarification to language, update timelines and list of collaborators |

## 4 Milestones

**Table 1.** Milestones.

| Milestone | Anticipated date of completion |
| --- | --- |
| Finalize study objectives and scope of work | February 2024 |
| Compile a list of risk factors for adjustment | May 2024 |
| Complete data preparation and statistical analysis | June 2024 |
| Discussion of results with collaborators | July 2024 |
| Manuscript draft and revisions | August 2024 |
| Submission to scientific journal | September 2024 |

## 5 Rationale and background

### What is known about the condition

Clinical trials are often criticized for lack of generalizability to real-world patient populations. This is for various reasons. Clinical trials often have restrictive eligibility criteria that exclude certain patient groups with disease, such as pregnant individuals or those with poor performance status. Additionally, some racial, socioeconomic and ethnic groups have been historically underrepresented in clinical trials. Furthermore, concomitant or subsequent therapies received in clinical trials in the US may differ from standards of care in routine care within the US as well as in other countries. For example, immunotherapy for aNSCLC is more commonly administered in the US than in Germany or Canada. It has been observed that patient outcomes are often better in clinical trials than in real-world studies (see efficacy-effectiveness gap).

Using statistical adjustment, it may be possible to model differences in patient, disease and treatment characteristics between trial and real-world populations to estimate real-world outcomes using data from a clinical trial. Consider the following scenarios:

a. Participants in clinical trial A were selected (possibly non-randomly) from the real-world population A’. We can therefore test the **<u>generalizability</u>** of patient outcomes in A to A’ if all patient subgroups in A’ were captured in A (i.e, A is a subset of A’).
b. Participants in clinical trial A were selected (possibly non-randomly) from the real-world population A’ but certain key patient groups in A’ were excluded from the trial. A’ is partly external to A and therefore we must test if we can **transport** from A to A’.
c. Participants in clinical trial A were selected from the real-world population A’ which is distinct from the real-world population B’ where prescribing decisions need to be made. A’ is partly external to A and thus we must test if we can **<u>transport</u>** from A to B’.
d. In this study, we aim to test scenarios b and c above using the US-based Lung-MAP sub-study S1400I (referred to as “Lung-MAP trial” herein) as a case study. Formally, our goal is to evaluate whether adjustment of patient survival in the Lung-MAP trial for important prognostic factors between patients in the Lung-MAP trial and real-world patients will recapitulate real-world patient survival.

### Note about terminology

A study sample, such as participants in a clinical trial, is selected from a larger target population of real-world patients. The following are interrelated concepts when discussing the appropriateness of results generated from the study sample in being applied to the target population.

External validity: Is an overarching concept that refers to how well results from the study sample apply, i.e. generalize or transport, to the target population. External validity contrasts with internal validity, which is whether the results are valid within the study sample. External validity can only be discussed if results are internally valid. Clinical trials aim to maximize internal validity through study design (e.g., through randomization).

Efficacy-effectiveness gap: Refers to the observation that patient outcomes or performance of therapies within clinical trials is often better than in routine care. This gap is a manifestation of limited external validity of results from clinical trials.

Generalizability: Refers to external validity when the study sample is a subset of the target population.

Transportability: Refers to external validity when the study sample is partly external to the target population. For example, this may refer to a clinical trial performed in the United States and a target population for the same indication in Germany. Because there are no German patients enrolled in the clinical trial, we discuss the transportability, rather than the generalizability, of results to Germany.

#### Squamous cell aNSCLC and Lung-MAP S1400I

Squamous cell lung carcinoma is a histological subtype of NSCLC that originates in the squamous cells lining the airways of the lungs. Historically, 25-30% of all cases of NSCLC are squamous cell carcinoma although these percentages can vary regionally and may change over time due to factors such as changes in smoking patterns. Compared to other NSCLC subtypes, such as adenocarcinoma, the presence of actionable genetic variants is less common and there are fewer targeted therapies available for squamous cell aNSCLC.

##### What is known about the exposure of interest

Patients diagnosed with squamous aNSCLC often receive Platinum-based chemotherapy regimens as first-line therapy. Following progression, patients will most often receive therapy with immune checkpoint inhibitors that target either the PD-(L)1 pathway, such as nivolumab and atezolizumab, or CTLA-4, such as ipilimumab. The Lung-MAP trial NCT02785952 compared overall survival in United States patients with recurrent stage IV squamous NSCLC randomized to receive either nivolumab monotherapy or nivolumab + ipilimumab combination therapy and found no significant difference in mortality rates between these groups.

- Note: Since FDA approval in October 2018 (<u>link</u>), combination therapy with pembrolizumab + chemotherapy has gradually replaced chemotherapy as first-line systemic treatment for patients with squamous aNSCLC. The NCT02785952 study was performed between 2016-2018 and the real-world studies between 2015-2020. For the NCT02785952 population, prior exposure to PD-L1 therapies was an exclusion criterion.

##### Gaps in knowledge

The transportability of the results from this trial, of both survival over time and relative effect measures (hazard ratio and risk difference) comparing second-line nivolumab to nivolumab + ipilimumab, to the real-world population in the United States, and in other countries, is unknown.

##### What is the expected contribution of this study?

This study has 3 goals:

1. The <u>primary goal</u> of this study is to assess the real-world generalizability of the findings of NCT02785952 to the US population, if possible, as well its transportability to real-world populations in the US, England, France, Japan and Germany. Because only second-line nivolumab is approved for use in these patients, we can only benchmark the generalizability/transportability of overall survival on second-line nivolumab monotherapy.
2. A secondary goal of this study is to estimate the relative effect of initiating nivolumab versus nivolumab + ipilimumab (risk difference and hazard ratio) after adjustment for risk factors of survival and effect measure modifiers. This will be a exploratory analysis to demonstrate transportability of relative effect measures rather than a benchmarking analysis because nivolumab + ipilimumab is not approved for second-line use for this indication and therefore no real-world data for it is available.
3. Another secondary goal of this study is to compare the reliability and speed of eliciting risk factors for adjustment from commonly available trained large language models (ChatGPT or GPT-3/4) versus those identified using subject matter expertise through a combination of manual search of the literature and input from oncologists.

### Justification for selection of the NCT02785952 trial for this study

For this demonstration study, we had access to data for trials available on Project Data Sphere (https://www.projectdatasphere.org/). To select a suitable trial, we identified all trials on Project Data Sphere that met the following criteria:

| Criterion for selection | Rationale | Number of studies available |
| --- | --- | --- |
| Individual-level data available on Project Data Sphere | Individual-level data is most conducive for parameter estimation for transportability analysis | 245 |
| Lung cancer tumour type | The authors of this study have considerable subject matter expertise in the advanced/metastatic lung cancer setting. Lung cancer also happens to be one of the most common types of cancer and a common target for therapeutic research. | 32 |
| Randomized phase III study designed to test efficacy | To avoid the issue of confounding and have reasonably large sample sizes for analysis | 24 |
| Testing an immunotherapy regimen that is commonly administered globally | For transportability analysis, we require real-world studies that have published data for patient characteristics and overall survival for the same intervention as the clinical trial. Immunotherapy was chosen because it is rapidly replacing/augmenting chemotherapy regimens and often prescribed broadly regardless of biomarker expression or genetic testing and therefore to be of broad interest. Their approval is also quite recent (post-2015) and therefore it is more likely that we will be examining recent and contemporaneous periods that are more relevant to a present day audience. | 1 |

## 6 Research question and objectives

**Table 2.**
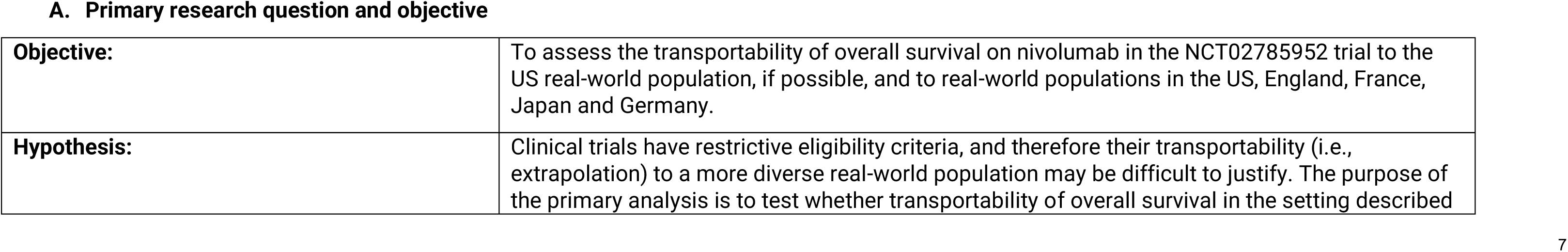
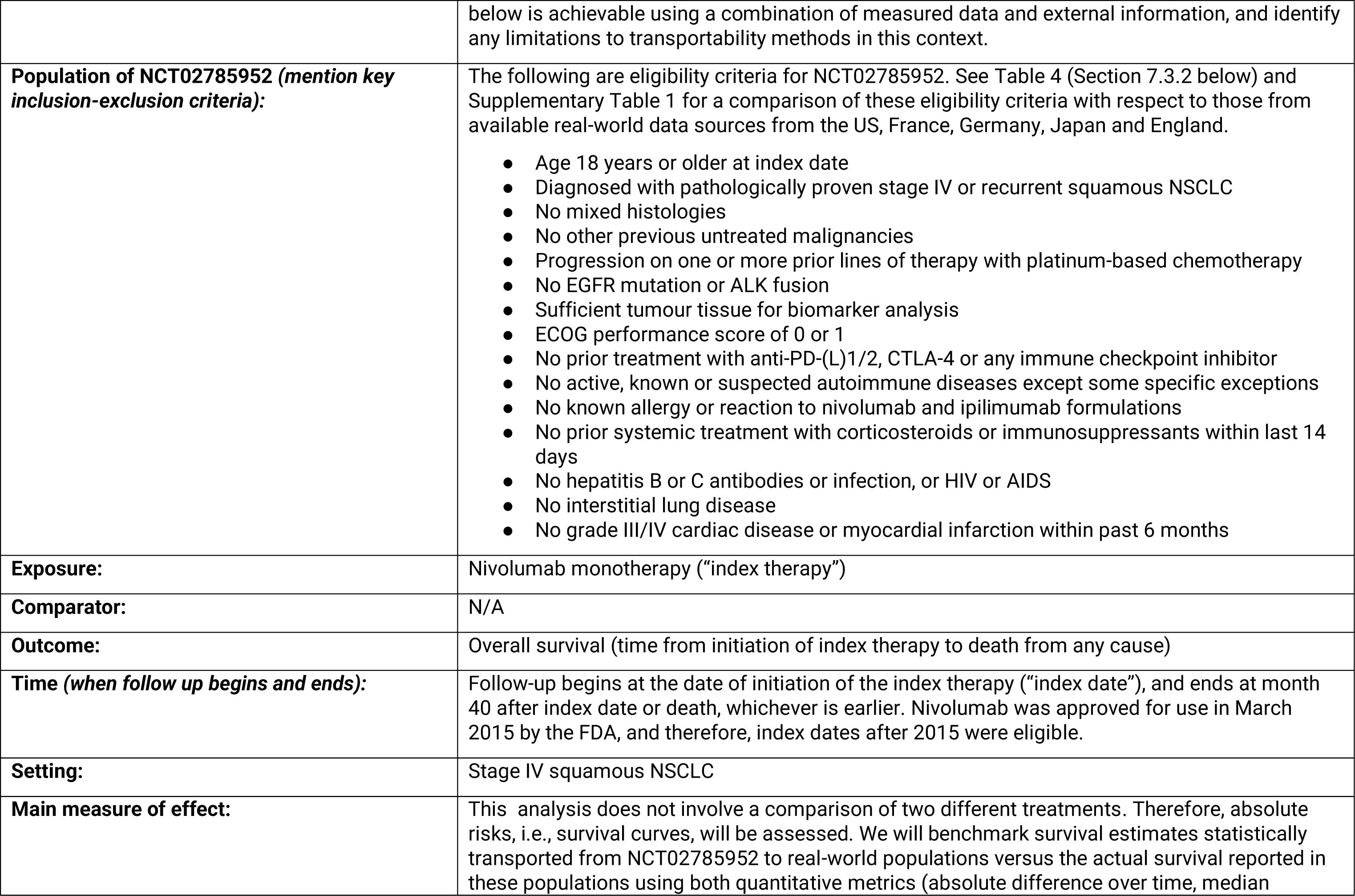
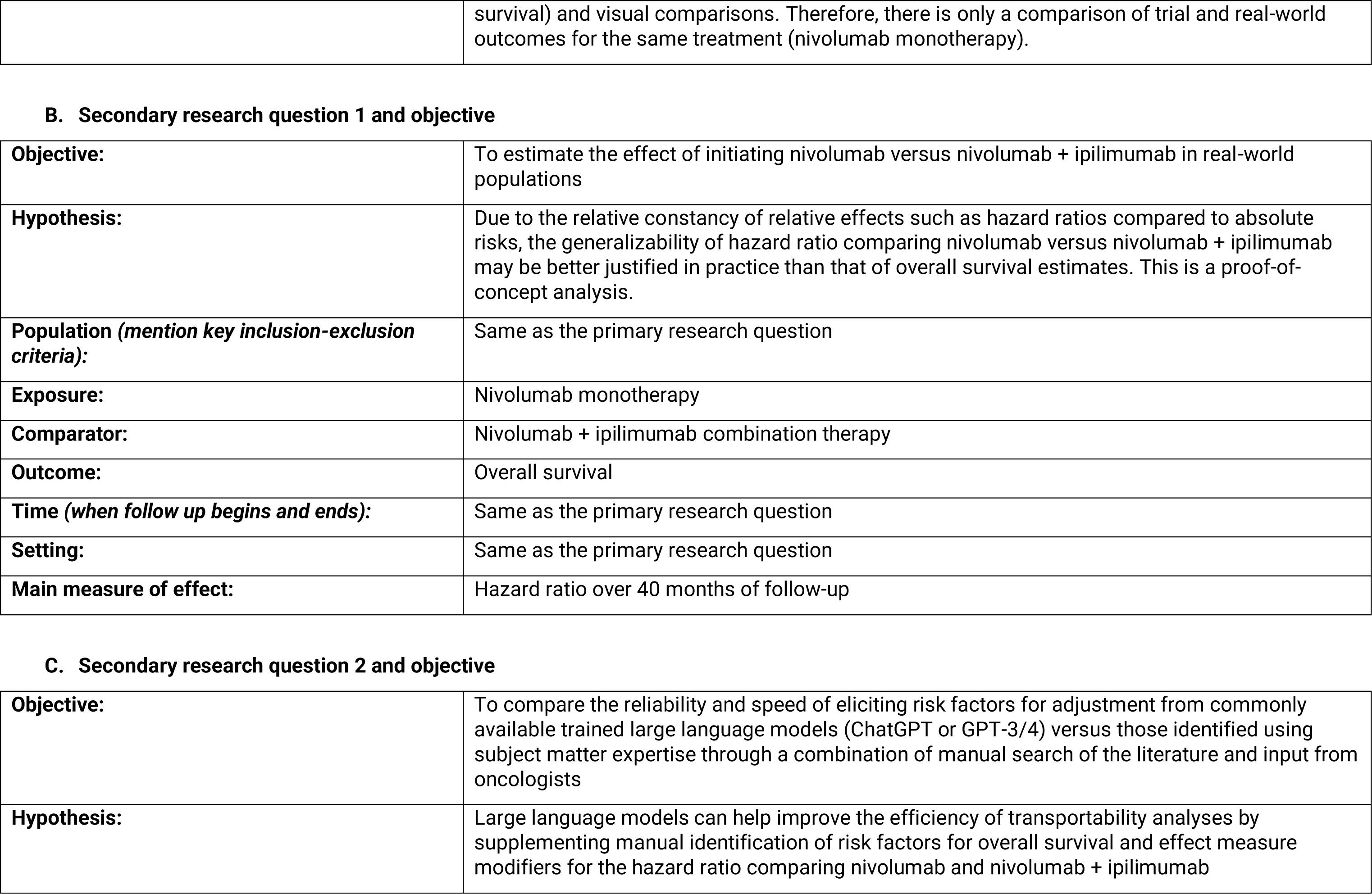
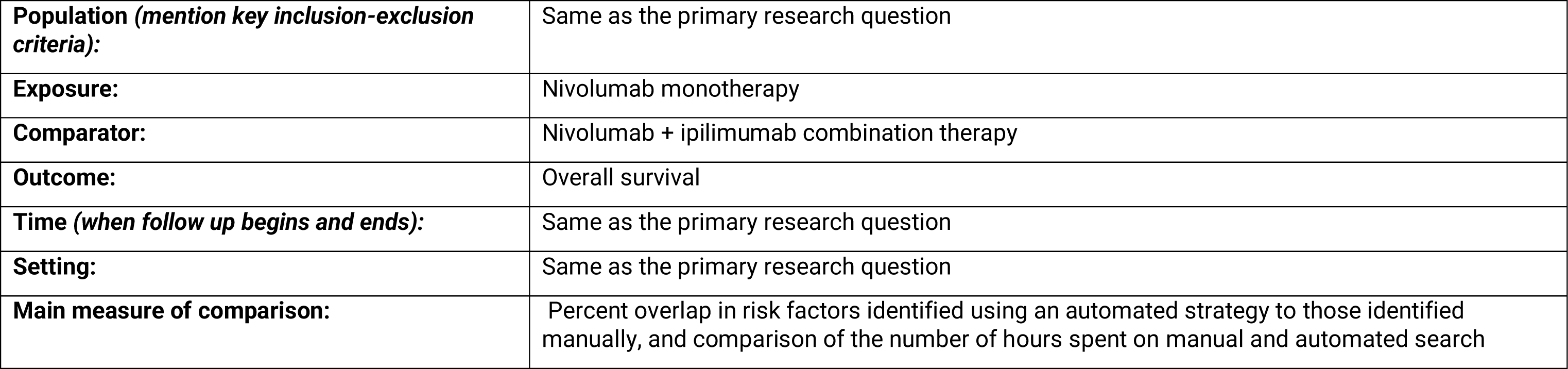
Primary and secondary research questions and objective.

A. Primary research question and objective
|  |  |
| --- | --- |
| **Objective:** | To assess the transportability of overall survival on nivolumab in the NCT02785952 trial to the US real-world population, if possible, and to real-world populations in the US, England, France, Japan and Germany. |
| **Hypothesis:** | Clinical trials have restrictive eligibility criteria, and therefore their transportability (i.e., extrapolation) to a more diverse real-world population may be difficult to justify. The purpose of the primary analysis is to test whether transportability of overall survival in the setting described |
|  | below is achievable using a combination of measured data and external information, and identify any limitations to transportability methods in this context. |
| **Population of NCT02785952 (mention key inclusion-exclusion criteria):** | The following are eligibility criteria for NCT02785952. See Table 4 (Section 7.3.2 below) and Supplementary Table 1 for a comparison of these eligibility criteria with respect to those from available real-world data sources from the US, France, Germany, Japan and England.<br>• Age 18 years or older at index date<br>• Diagnosed with pathologically proven stage IV or recurrent squamous NSCLC<br>• No mixed histologies<br>• No other previous untreated malignancies<br>• Progression on one or more prior lines of therapy with platinum-based chemotherapy<br>• No EGFR mutation or ALK fusion<br>• Sufficient tumour tissue for biomarker analysis<br>• ECOG performance score of 0 or 1<br>• No prior treatment with anti-PD-(L)1/2, CTLA-4 or any immune checkpoint inhibitor<br>• No active, known or suspected autoimmune diseases except some specific exceptions<br>• No known allergy or reaction to nivolumab and ipilimumab formulations<br>• No prior systemic treatment with corticosteroids or immunosuppressants within last 14 days<br>• No hepatitis B or C antibodies or infection, or HIV or AIDS<br>• No interstitial lung disease<br>• No grade III/IV cardiac disease or myocardial infarction within past 6 months |
| **Exposure:** | Nivolumab monotherapy (“index therapy”) |
| **Comparator:** | N/A |
| **Outcome:** | Overall survival (time from initiation of index therapy to death from any cause) |
| **Time (when follow up begins and ends):** | Follow-up begins at the date of initiation of the index therapy (“index date”), and ends at month 40 after index date or death, whichever is earlier. Nivolumab was approved for use in March 2015 by the FDA, and therefore, index dates after 2015 were eligible. |
| **Setting:** | Stage IV squamous NSCLC |
| **Main measure of effect:** | This analysis does not involve a comparison of two different treatments. Therefore, absolute risks, i.e., survival curves, will be assessed. We will benchmark survival estimates statistically transported from NCT02785952 to real-world populations versus the actual survival reported in these populations using both quantitative metrics (absolute difference over time, median |
|  | survival) and visual comparisons. Therefore, there is only a comparison of trial and real-world outcomes for the same treatment (nivolumab monotherapy). |
| B. Secondary research question 1 and objective |  |
| **Objective:** | To estimate the effect of initiating nivolumab versus nivolumab + ipilimumab in real-world populations |
| **Hypothesis:** | Due to the relative constancy of relative effects such as hazard ratios compared to absolute risks, the generalizability of hazard ratio comparing nivolumab versus nivolumab + ipilimumab may be better justified in practice than that of overall survival estimates. This is a proof-of-concept analysis. |
| **Population (*mention key inclusion-exclusion criteria*):** | Same as the primary research question |
| **Exposure:** | Nivolumab monotherapy |
| **Comparator:** | Nivolumab + ipilimumab combination therapy |
| **Outcome:** | Overall survival |
| **Time (*when follow up begins and ends*):** | Same as the primary research question |
| **Setting:** | Same as the primary research question |
| **Main measure of effect:** | Hazard ratio over 40 months of follow-up |
| C. Secondary research question 2 and objective |  |
| **Objective:** | To compare the reliability and speed of eliciting risk factors for adjustment from commonly available trained large language models (ChatGPT or GPT-3/4) versus those identified using subject matter expertise through a combination of manual search of the literature and input from oncologists |
| **Hypothesis:** | Large language models can help improve the efficiency of transportability analyses by supplementing manual identification of risk factors for overall survival and effect measure modifiers for the hazard ratio comparing nivolumab and nivolumab + ipilimumab |
| **Population (*mention key inclusion-exclusion criteria*):** | Same as the primary research question |
| **Exposure:** | Nivolumab monotherapy |
| **Comparator:** | Nivolumab + ipilimumab combination therapy |
| **Outcome:** | Overall survival |
| **Time (*when follow up begins and ends*):** | Same as the primary research question |
| **Setting:** | Same as the primary research question |
| **Main measure of comparison:** | Percent overlap in risk factors identified using an automated strategy to those identified manually, and comparison of the number of hours spent on manual and automated search |

### Potential use cases for this research and target audience

The target audience for this research includes public health researchers interested in understanding the efficacy-effectiveness gap, health technology assessment bodies interested in transportability of results from US trials or real-world studies to local contexts, and researchers interested in global health or analytical methods. The potential use cases for this research include the following:

1. Regulators are increasingly interested in diversity in clinical trials. Due to logistical issues, however, it may be challenging to diversify explicit and implicit selection criteria in clinical trials or alter how trials are implemented. This study hopes to identify factors, as well as methodological opportunities and limitations, in using trial data to estimate treatment effects or patient outcomes in a diverse population.
2. Decision-making in the clinic using results from trials after standardizing them to the population that is most likely to be observed in the clinic.
3. External control arms selected from real-world data sometimes produce outcomes that are worse than would be observed with a concurrent control in a clinical trial, leading to an exaggerated benefit for the experimental therapy. This problem may be due to unemulatable eligibility criteria and may persist even after adjustment for measured confounders. Patient outcomes for either the trial arm or the external control arm may be adjusted using statistical adjustment methods to better represent the target population.

## 7 Research methods

### 7.1. Study design

**Research design (e.g. cohort, case-control, etc.):** Indirect treatment comparison

**Rationale for study design choice:** For assessing transportability of patient risks from a clinical trial, a cohort study is most appropriate.

### 7.2. Study design diagram

The specifics may differ for different data sources.

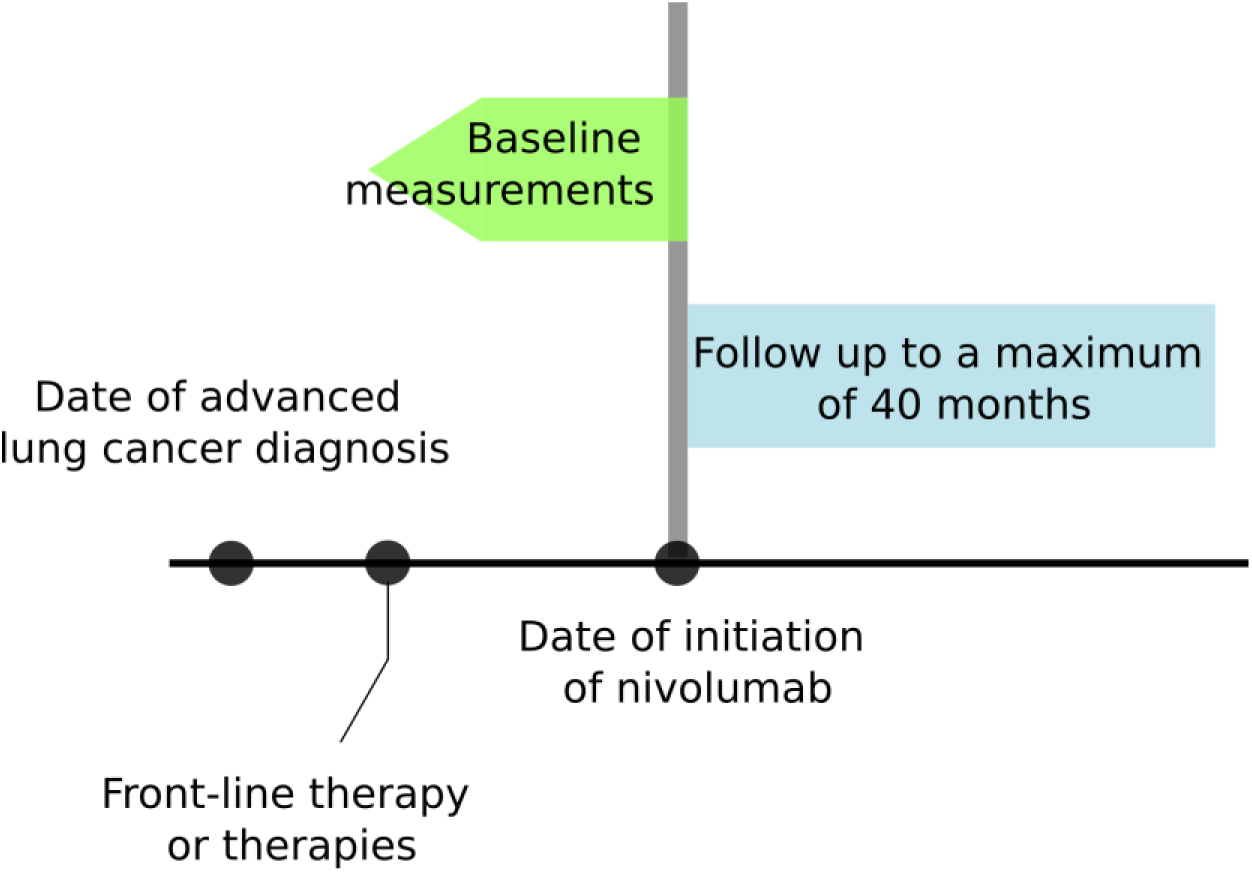

Except for the English study (IO-Optimise), which uses the date of diagnosis as index date, all other real-world studies use the date of initiation of nivolumab as the index date. Given that this is likely to be an important difference that affects transportability, we hypothesize that the English study specifically will act as a negative control and help understand the limitations of statistical analysis for transportability estimation.

As a part of the study, the study design across the different studies will be tabulated in a form similar to that used for target trial emulation.

### 7.3. Setting

#### 7.3.1 Context and rationale for definition of time 0 (and other primary time anchors) for entry to the study population

The index date is the date of treatment assignment in NCT02785952.

For the IO-Optimise study, time zero was the time of initial diagnosis, which is quite different.

**Table 3.**
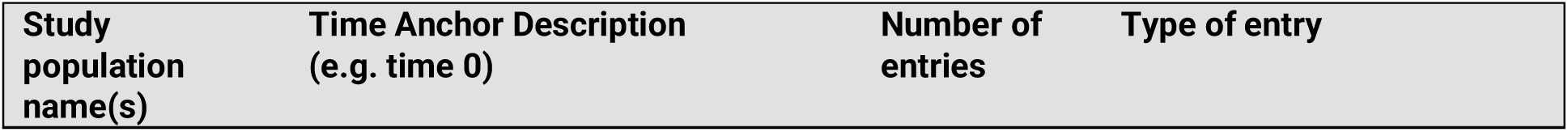
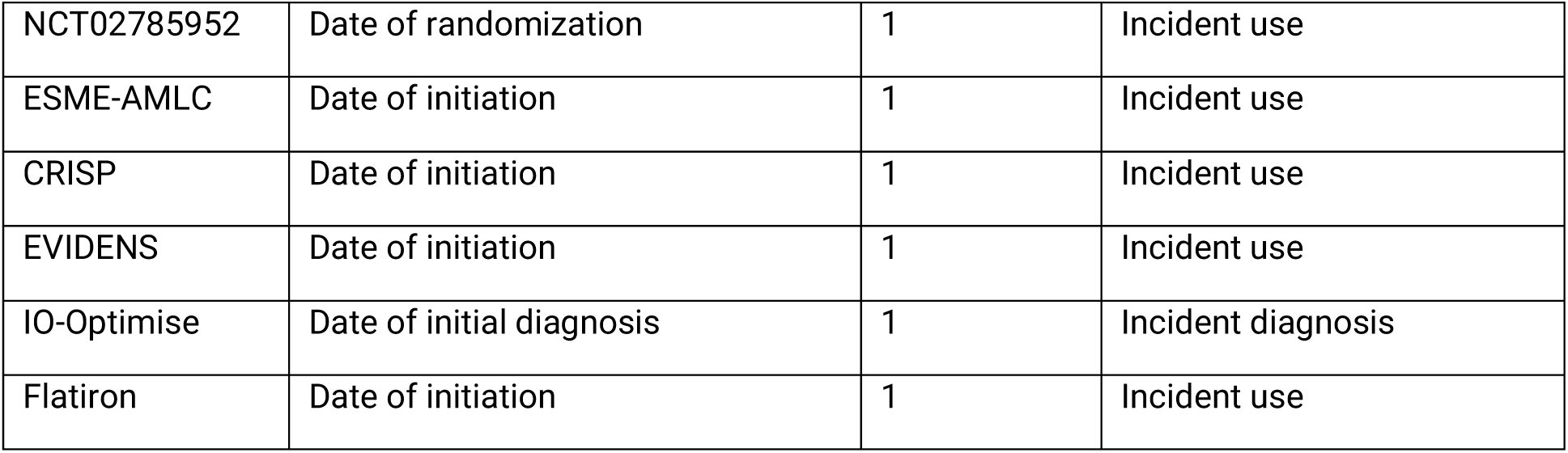
Operational Definition of Time 0 (index date) and other primary time anchors.

#### 7.3.2 Context and rationale for study eligibility criteria

Study inclusion criteria differ between the source data from the NCT02785952 trial and those for target real-world populations from England, Germany, Japan and France. The eligibility criteria for both source and target data is fixed and can not be altered. A comparison of the eligibility criteria, those reported, is provided here:

**Table 4.**
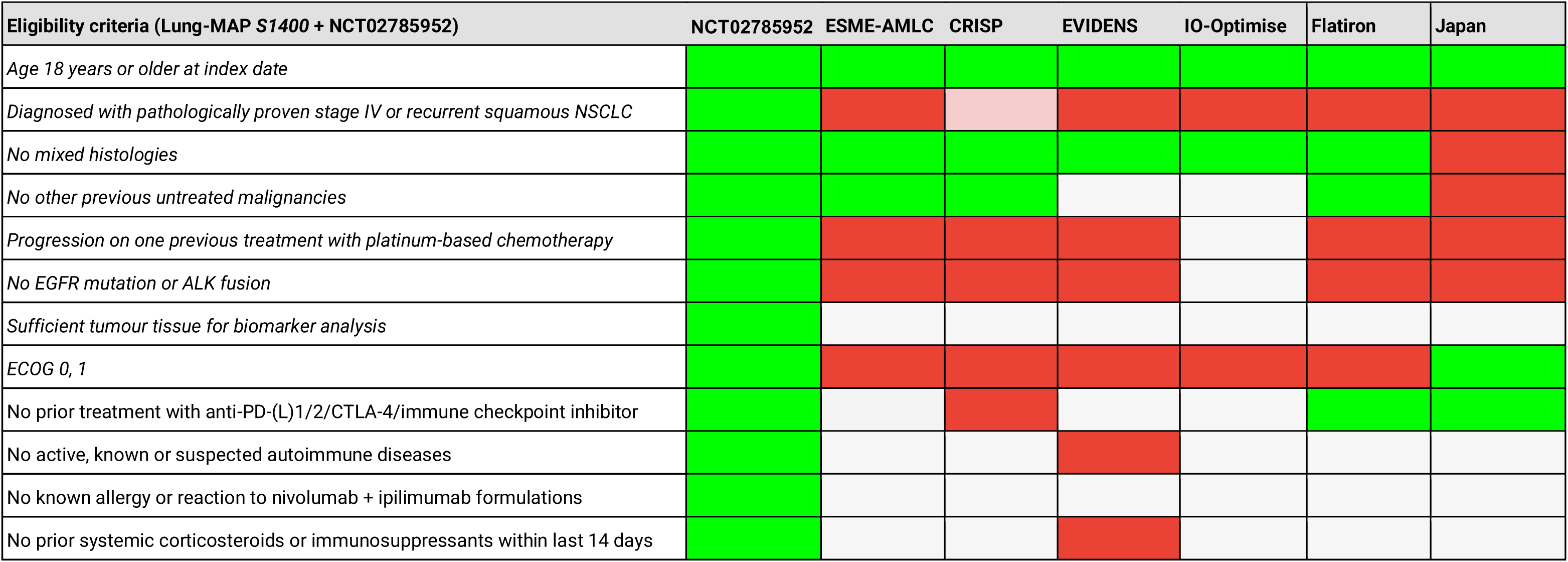
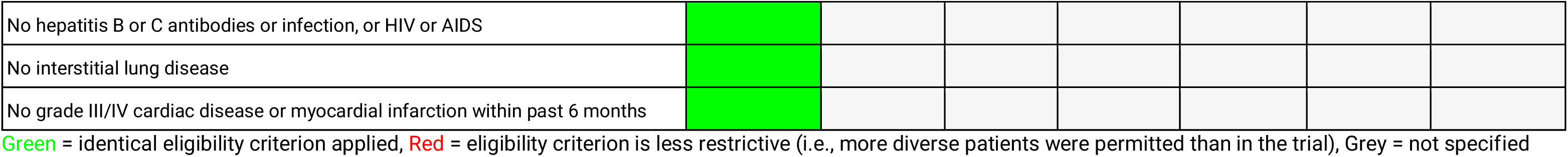
Eligibility criteria across data sources.

### 7.4 Variables

Context, rationale, and operation definition of exposures, outcomes, and other variables including measured risk factors, comorbidities, co-medications, potential confounding variables and effect modifiers, etc. are specified in this section.

#### 7.4.1 Context and rationale for exposure(s) of interest

The treatment groups of interest are the same as those in NCT02785952. For real-world studies, we make the concession that any dose and frequency compatible with observed data is permitted, even if not reported. This is defensible because FDA has approved both 3 mg/kg nivolumab and 240 mg fixed dose based on pharmacokinetic data that suggested no difference between these regimens, and patient outcomes do not seem to differ greatly between these two treatment versions in practice [https://www.ncbi.nlm.nih.gov/pmc/articles/PMC7004547/]. Both regimens are approved in Europe (cf. EVIDENS study).

Treatment regimens in NCT02785952:

- Nivolumab administered intravenously at a dose of 3 mg/kg every 14 days
- Nivolumab administered intravenously at a dose of 3 mg/kg every 14 days + ipilimumab given at 1 mg/kg on day 1 of every third cycle

#### 7.4.2 Context and rationale for outcome(s) of interest

Overall survival defined as time from index date to death from any cause is the primary and sole outcome of interest. Overall survival is the most important clinical outcome in aNSCLC.

**Table 5.** Operational Definitions of Outcome.

| Outcome name | Details | Primary outcome ? | Type of outcome | Washout window | Care Settings<sup>1</sup> | Code Type<sup>2</sup> | Diagnosis Position<sup>3</sup> | Applied to study populations: | Measurement characteristics/ validation | Source of algorithm |
| --- | --- | --- | --- | --- | --- | --- | --- | --- | --- | --- |
| Overall survival | Time from index date to death | Yes | Time-to-event | None | N/A | N/A | N/A | Target populations specified in Table 2 | Unless specified, assumed to be reliable | N/A |
|  | from any cause |  |  |  |  |  |  |  | ESME-AMLC:<br>not cross-checked with national registry |  |

#### 7.4.3 Context and rationale for follow up

The maximum length of follow-up in the NCT02785952 data cut available to us is approximately 40 months. Therefore, 40 months from index date/time zero is the maximum follow-up we can assess for transportability using NCT02785952. For the English study, where index date is date of diagnosis rather than date of nivolumab initiation, the 40 month follow-up begins at date of diagnosis.

**Table 6.** Operational Definitions of Follow Up. Except for the English study (IO-Optimise), which uses the date of diagnosis as index date, all other real-world studies use the date of initiation of nivolumab as the index date. NCT02785952 uses the date of randomization, i.e., assignment, as the index date.

|  |  |  |
| --- | --- | --- |
| **Follow up start** | Day 1 |  |
| **Follow up end<sup>1</sup>** | Select all that apply | **Specify** |
| **Date of outcome** | Yes | Outcome is death for any reason |
| **Date of death** |  |  |
| **End of observation in data** | Yes | The administrative end of follow-up is different for different real-world databases |
| **Day X following index date**<br>(specify day) | Yes | 40 months from index date |
| **End of study period**<br>(specify date) | No | N/A |
| **End of exposure**<br>(specify operational details,<br>e.g. stockpiling algorithm, grace period) | No | N/A |
| **Date of add to/switch from exposure**<br>(specify algorithm) | No | N/A |
| **Other date (specify)** | None | N/A |

<sup>1</sup> Follow up ends at the first occurrence of any of the selected criteria that end follow up.

#### 7.4.4 Context and rationale for covariates (confounding variables and effect modifiers, e.g. risk factors, comorbidities, comedications)

For transportability of absolute risks using the outcome model as in this study, an assumption for unbiased estimation is adjustment for all risk factors for the outcome are included in the outcome model that differ between the trial and real-world cohorts.

**Table 7.** Operational Definitions of Covariates. The following is a tentative list of risk factors for overall survival we attempt to adjust for. Variables that are measured in NCT02785952 data available for this study are marked with an asterisk (*). A comparison of available covariates and their values from NCT02785952 and published studies can be found in Appendix table 1. The operational definitions of covariates from published studies is not directly available.

| Characteristic | Details | Type of variable | Assessment window |
| --- | --- | --- | --- |
| Age* | Age (in years) at randomization | Continuous | Screening variables were assessed within 14 days of registration in NCT02785952 |
| Sex* | Sex | Dichotomous (male or female) |  |
| ECOG score* | ECOG performance score (0-4) | Ordinal |  |
| Liver metastases* | Presence of liver metastases at baseline | Dichotomous (present or absent) |  |
| Brain metastases* | Presence of brain metastases at baseline | Dichotomous (present or absent) |  |
| Smoking history* | History of smoking | Dichotomous (ever or never) |  |
| Time since diagnosis | Time in months since advanced diagnosis | Continuous |  |
| Number of prior lines of therapy* | Number of prior lines of systemic therapy at baseline | Dichotomous (1 or 2+) |  |
| EGFR mutations | Presence of EGFR variants | Dichotomous (present or absent) |  |
| PD-L1 expression | Expression of PD-L1 | Dichotomous (present or absent) |  |

\* = Variables available in NCT02785952 for which individual-level data

### 7.5 Data analysis

#### 7.5.1 Context and rationale for analysis plan

The inferential data analysis plan is described conceptually, and the context or rationale for the choices are provided in this section.

For the primary analysis of transportability of survival estimates, the parametric G-formula will be used. The reason for this choice, rather than a model using inverse probability weighting, is twofold. First, because real-world studies include more diverse populations than the clinical trial population due to less stringent eligibility criteria, we rely on extrapolation from the outcome model and assess whether such extrapolation is sensible and reliable. This can not be done using an inverse weighting method. Second, due to the presence of unmeasured risk factors, we are planning to use external information from published multivariable analyses to inform sensitivity analyses. We anticipate that such external information will be available in the form of coefficients of multivariable analyses in published articles. G-computation also allows us to simulate counterfactual risks for unmeasured time-varying risk factors related to differences in subsequent therapies between countries as a time-varying prognostic variable (see Ramagopalan et al. 2023 for details). However, an inverse probability model will be reported, sufficiently high effective sample sizes are recovered, an inverse probability weighted analysis including only for baseline risk factors will also be performed.

For the secondary analysis, relative risks in the form of hazard ratios and risk difference will be calculated using contrasts based on transported survival curves informed by the primary analysis.

**Table 8.** Primary, secondary, and subgroup analysis specification.

**A. Primary analysis**
|  |  |
| --- | --- |
| **Hypothesis:** | The combination of measured data and external information can facilitate the successful extrapolation of overall survival outcomes from clinical trials with restrictive eligibility criteria to a more diverse real-world population, and the primary analysis will reveal insights into the feasibility and limitations of transportability methods in this specific context. |
| **Exposure contrast:** | No contrasts. Only nivolumab monotherapy will be assessed. |
| **Outcome:** | Overall survival |
| **Analytic software:** | R version 4.3<br>Packages: survival, survminer, gfoRmula, copula (Package versions and any additional packages used for the analysis will be documented) |
| **Model(s):**<br>**(provide details or code)** | Anticipated primary model based on measured risk factors (the exact specification may change subject to clinical input during the project):<br>`death ~ age*sex*ecog*TRT + smoking_status + num_prior_txlines + brainmets + livermets + splines(time)`<br>An additional model augmented with unmeasured baseline and time-varying risk factors will be generated, which will also attempt to account for positivity violations (see section below on “Use of external data to handle positivity violations”). The proposed specification for this model is currently as follows, and is subject to change:<br>`death ~ age^2 + age*sex*ecog + race + num_prior_txlines + time_since_diag + time_since_diag^2 + race + smoker + splines(time) + brainmets + livermets + egfr + pdl1` |
| **Adjustment method** | ***Name method and provide relevant details, e.g. bivariate, multivariable, propensity score matching (specify matching algorithm ratio and caliper), propensity score weighting (specify weight formula, trimming, truncation), propensity score stratification (specify strata definition), other.*** |
|  | The outcome model specified above will be used to adjust for measured differences in baseline risk factors. Other specifications will be used for sensitivity analysis. |
| **Missing data methods** | ***Name method and provide relevant details, e.g. missing indicators, complete case, last value carried forward, multiple imputation (specify model/variables), other.*** |
|  | There are no missing values in the NCT02785952 dataset. There is missing data for individual measured variables in published real-world studies (see Appendix table 1). For example, the prevalence of ECOG score of 0 was only reported within the subset of patients of patients with non-missing ECOG scores. We assume that the marginal distribution of these variables is similar to the reported summary-level distributions and use the Copula method for generating 1000 imputed datasets using the covariance matrix estimated from trial data (which contains no missing data) and summary-level data from published real-world studies. |
| **Subgroup Analyses** | ***List all subgroups*** |
|  | None |

##### Benchmarking analysis

There is no single benchmarking metric that can comprehensively quantify the success or feasibility of transportability. To be compatible with regulatory, health technology assessment and clinical use cases, we propose the following benchmarking metrics comparing estimated versus observed survival:

- Median survival time
- Mean survival probability at the end of follow-up (40 months)
- Area under survival curves (i.e., restricted mean survival) from time zero until end of follow-up
- Clinically significant differences based on evaluation by oncologist collaborators

##### Use of external data to handle positivity violations

A formal description of transportability analysis with G-computation with structural positivity violations augmented with external information can be found in Zivich et al. (Epidemiology, 2024). We describe the proposed analysis based on Zivich et al. here in plain text as it applies to this study:

1. We specify a model for the outcome “Mortality” as a function of both measured and unmeasured baseline and time-varying risk factors
2. Some of the parameters of this model (parameters A) representing the can be estimated using trial data. These parameters represent (informally) the association of measured risk factors with survival within this population.
3. The remaining parameters (parameters B) need to be estimated from external data (e.g., Cox regression coefficients from published literature). If no data is available, an educated clinical guess will be used. Parameters B will be specified with a distribution. [Technical note: Details about marginal versus conditional values for parameters will be justified based on available information from the trial and external information where necessary].
4. The probability of the outcome is equal to the model with estimated parameters A + B after applying the appropriate link function.

This procedure for estimation of confidence intervals is conceptually similar to a quantitative probabilistic bias analysis for unmeasured variables.

##### Sensitivity analysis

Sensitivity analyses will be performed for specification of the parametric outcome model. This is done because the sample size in the trial (n=127 for the nivolumab monotherapy arm) will limit the complexity of the outcome model that we can feasibly estimate parameters for, potentially leading to risk of model misspecification with an overly simple model. Sensitivity analyses using a variety of combinations of interaction and higher-order terms (possibly randomly chosen) will be used to document the sensitivity of the predicted risks within the trial to the outcome model specification.

B. Secondary Analysis 1
|  |  |
| --- | --- |
| **Hypothesis:** | Hazard ratios may offer more practicable generalizability than overall survival estimates when comparing nivolumab versus nivolumab + ipilimumab, as demonstrated in this proof-of-concept analysis. |
| **Exposure contrast:** | Nivolumab monotherapy versus nivolumab + ipilimumab combination |
| **Outcome:** | Overall survival |
| **Analytic software:** | Same as primary analysis |
| **Model(s):**<br>(provide details or code) | Same as primary analysis |
| **Adjustment method** | *Name method and provide relevant details, e.g. bivariate, multivariable, propensity score matching (specify matching algorithm ratio and caliper), propensity score weighting (specify weight formula, trimming, truncation), propensity score stratification (specify strata definition), other.* |
|  | Same as primary analysis |
| **Missing data methods** | *Name method and provide relevant details, e.g. missing indicators, complete case, last value carried forward, multiple imputation (specify model/variables), other.* |
|  | Same as primary analysis |
| **Subgroup Analyses** | *List all subgroups* |
|  | Same as primary analysis |

Secondary analysis 2 is not a comparative analysis of exposures. To implement automated identification of risk factors for survival in lung cancer, they will be elicited using GPT-3/4 language models by a third party. These will then be compared to the primary analysis in speed (time taken to identify risk factors and published studies supporting these) and overlap in identified factors.

**Table 9.** Sensitivity analyses – rationale, strengths and limitations. The table below details the sensitivity analyses, the rationale for conducting them (in other words, stating what the investigator intends to learn from doing the sensitivity analysis), and any strengths or limitations of the sensitivity analysis relative to the primary analysis. Other sensitivity analyses will be performed on an ad hoc basis to account for uncertainties that arise during the analysis.

|  | What is being varied? How? | Why?<br>(What do you expect to learn?) | Strengths of the sensitivity analysis compared to the primary | Limitations of the sensitivity analysis compared to the primary |
| --- | --- | --- | --- | --- |
| Outcome model specification | Outcome model | Sensitivity of results to unmeasured risk factors | Complements the primary analysis by providing evidence of results to unmeasured data | Parameters to inform this sensitivity analysis are based on external information and may not be as reliable as measured data, or as informative. This is quantitative bias analysis. |

### 7.6 Data sources

#### 7.6.1 Context and rationale for data sources

##### Reason for selection

NCT02785952 was selected from Project Data Sphere as the study population. The criteria for selecting a suitable clinical trial dataset for this study were as follows: (i) a lung cancer indication, (ii) a phase III randomized trial with both treatment arms available, with sample size >100 and (iii) testing a non-chemotherapy regimen of clinical importance that is approved for use and commonly administered outside the United States. NCT02785952 was the only trial on Project Data Sphere that fit these criteria. aNSCLC was chosen because the study authors have substantial experience in this disease setting, and because it is a common indication for drug development, and therefore for regulatory approvals and health technology assessments.

Subsequently, corresponding real-world studies from the US, Germany, Japan, France and England were identified from the literature. Published studies must have been performed in patients with aNSCLC who initiated nivolumab monotherapy, and reported baseline characteristics and a Kaplan-Meier curve for overall survival in these patients for a squamous histological subtype, as in NCT02785952.

##### Strengths of data source(s)

Patient-level data is available for important risk factors and overall survival.

##### Limitations of data source(s)

The original ADaM/SDTM dataset is not available, and therefore we only have derived variables in some cases. No longitudinal data on risk factors or subsequent therapies is available.

##### Data source provenance/curation

Not available for the available dataset. However, the distribution of baseline characteristics and Kaplan-Meier estimates match those from the trial publication (not shown here).

**Table 10.** Metadata about data sources and software. Details of the individual-level patient data for NCT02785952 are shown below.

|  | **Trial data** |
| --- | --- |
| **Data Source(s):** | NCT02785952 |
| **Study Period:** | 2015-12-29 to 2019-12-19 based on https://www.clinicaltrials.gov/study/NCT02785952. |
| **Eligible Cohort Entry Period:** | Not known. The data available does not include this information. |
| **Data Version (or date of last update):** | Not known |
| **Data sampling/extraction criteria:** | N/A |
| **Type(s) of data:** | Patient-level data from a randomized clinical trial |
| **Data linkage:** | N/A |
| **Conversion to CDM\*:** | None |
| **Software for data management:** | None |

\*CDM = Common Data Model

### 7.7 Data management

Patient-level data from NCT02785952 has been provided by the trial sponsor in a deidentified format. A single copy of this dataset will be stored on a local password-protected computer. Programming code will be stored and backed up in a private and secured cloud repository.

### 7.8 Quality control

Data from published studies for real-world populations can not be manually verified by us with respect to data quality, reliability or validation. We assume that these steps were taken by the authors. For NCT02785952, we have a limited dataset provided by the trial sponsor. Although double programming will not be performed, steps will be taken to ensure that there are no programming errors for data processing and analysis that can affect the accuracy of the results, for example, through sanity checks and visualization of intermediate results/outputs in the analysis.

### 7.9 Study size and feasibility

Because this is an exploratory study, sample size calculations were not formally performed. However, results from a feasibility analysis not shown here indicate that there NCT02785952 has a large enough sample size to be able to estimate parameters of the outcome model whose signs (negative or positive coefficient) are consistent with prior expectations about the positive or negative correlation of risk factors with patient survival in aNSCLC.

#### Table 13. Power and sample size

Not applicable. This is an exploratory study.

## 8 Limitation of the methods

The following is a discussion of potential limitations of the study design, and analytic methods, including issues relating to confounding, bias, generalisability, and random error:

- Random error – NCT02785952 is a relatively small trial (125 + 127 patients) and therefore the results may have low precision. This limitation exists for any study with small samples and is a common concern in external control arm studies. 95% confidence intervals will be estimated, and any attempts to improve precision will be documented, with accompanying sensitivity analyses.
- Extrapolation beyond the study sample without reliable external information – As shown in Section 7.3.2, the target populations represented by the real-world studies used in this analysis have less restrictive eligibility criteria than NCT02785952. Therefore, we are relying on extrapolation based on the outcome model to these populations. We will attempt to use external information from published multivariable analyses in lung cancer to inform parameters for population characteristics represented in target datasets but not in NCT02785952, for example, the multivariable association of ECOG=2 or 3 with overall survival. However, the external information may not be within an identical target population, or there may be no data available for certain parameters, in which case educated guesses will need to be used.
- Differences in variable measurement across datasets – There are known differences between the datasets in measured variables. There is also the possibility that there are other unknown differences in variable recording and derivation across the datasets. Important differences will be flagged by comparison of baseline characteristics across studies for these variables for sensitivity analysis or quantitative bias analysis.
- Misspecification of the outcome model – Outcome models are susceptible to misspecification. Although we can not rule out model misspecification, we will test the sensitivity of model parameters and the predicted outcome under natural course using multiple specifications of the outcome model, including those with higher-order and interaction terms. Planned sensitivity analyses for these are documented in section 7.5.
- However, not all risk factors for survival are measured across all data sources. Moreover, it is impossible to adjust for all risk factors even if they were measured and impossible to know for sure that we have adjusted for a sufficient number of risk factors beyond which prognostic association of any additional variables with survival is small. However, we will attempt to justify the choice of risk factors in our study and provide bounds, if possible, on sensitivity of results to unmeasured factors.
- Lack of individual-level data for the target populations, and lack of longitudinal data for the trial and real-world sources on subsequent therapies and disease progression
- Because we are testing a single indication in this study using the Lung-MAP study, our results may not generalize to other indications. However, our study will aim to provide a guide for future research in other indications and therapies.

## 9 Protection of human subjects

All data was de-identified.

## 10 Reporting of adverse events

Not applicable

## Data Availability

Data can be requested through Project Data Sphere's Data Sharing Platform.

## 12. Appendices

**Appendix table 1.**
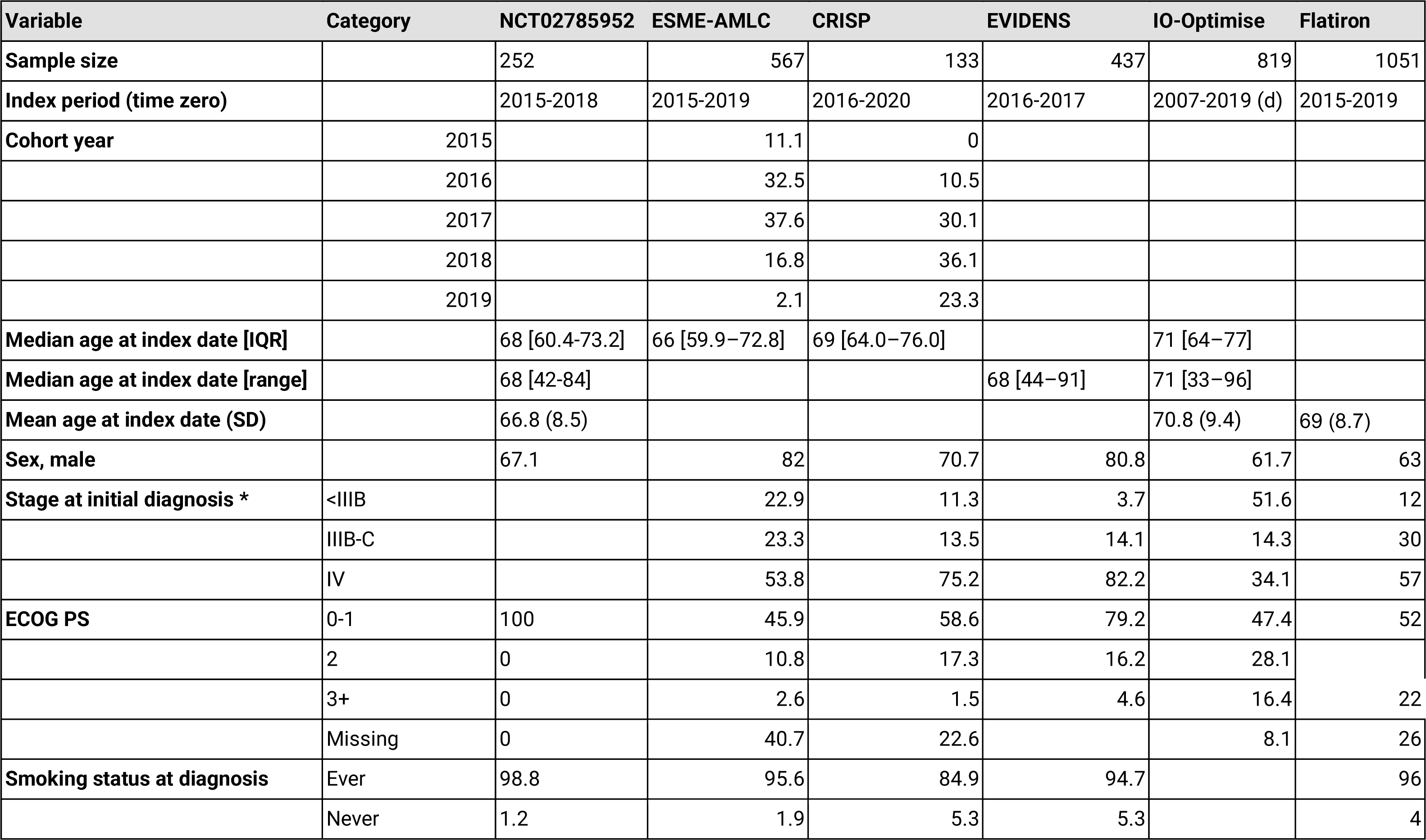
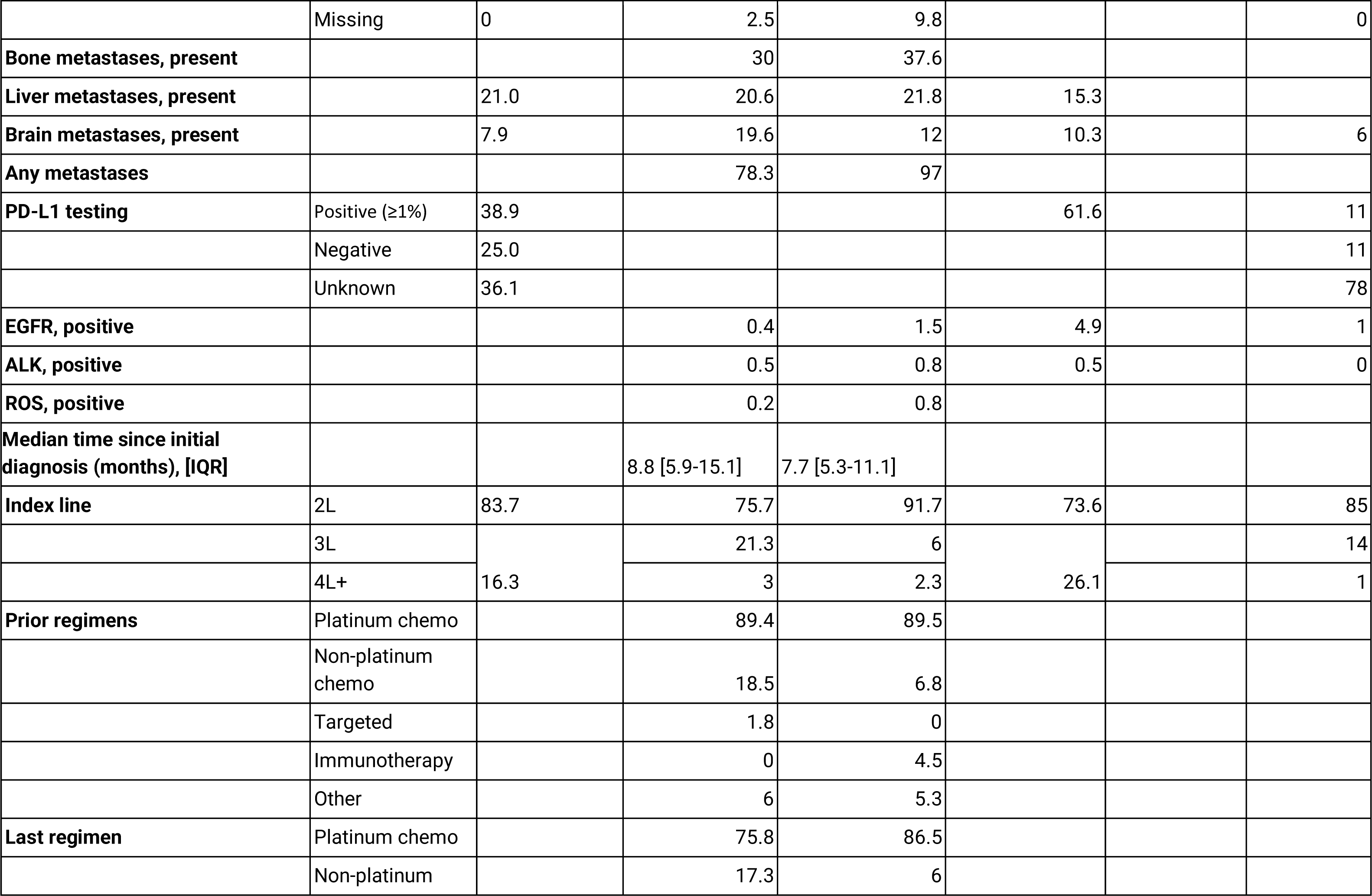
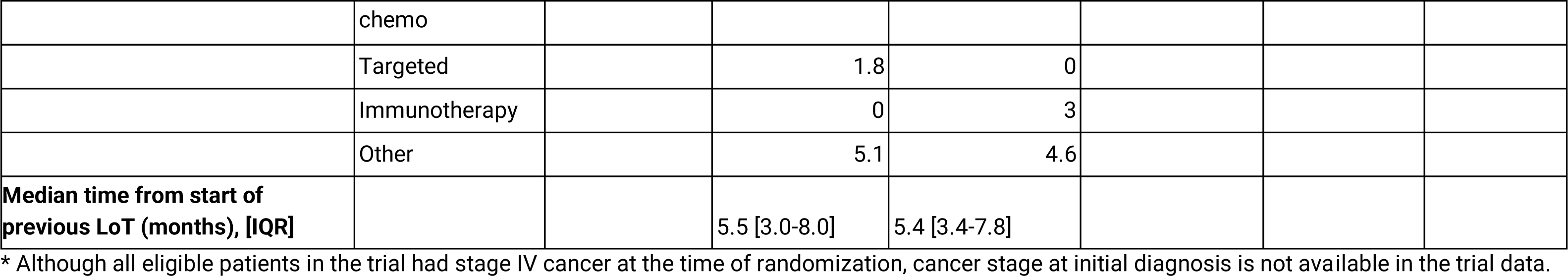
Distribution of baseline covariates across datasets in this study. Unfilled categories imply lack of reported or measured information.

## Notes

**Conflict of interest** None

### Competing Interest Statement

The authors have declared no competing interest.

### Funding Statement

This study did not receive any funding

### Author Declarations

This work was performed using data from the NCTN Data Archive of the National Cancer Institute's (NCI's) National Clinical Trials Network (NCTN). Data were originally collected from clinical trial NCT number NCT02785952. Data can be accessed through Project Data Sphere's Data Sharing Platform.

### Summary of Updates

Added co-authors to the author list on medRxiv. No change to the protocol.

